# Analytical and clinical validation of a genome sequencing-based comprehensive rare disease genomic profiling test

**DOI:** 10.1101/2024.10.19.24315813

**Authors:** Shruthi Sriram, Sang Yeon Lee, Stephanie Ferguson, June-Young Koh, Jonathan Kyle Wallace, Jeonghoon Lee, Jung-Ah Kim, Yoonsuh Lee, Brian Baek-Lok Oh, Won Chul Lee, Sangmoon Lee, Erin Connolly-Strong

## Abstract

This study evaluates the performance of the RareVision Whole Genome Sequencing (WGS) assay for comprehensive genomic profiling in rare genetic diseases. The analytical validation assessed the assay’s sensitivity and positive predictive values (PPV) for single nucleotide variants (SNVs), insertions/deletions (indels), and structural variants (SVs), revealing a sensitivity of 99.4% for SNVs and 98.7% for indels, with PPVs of 99.3% for SNVs and 98.7% for indels. Clinical validation involved benchmarking against established orthogonal methods, demonstrating high concordance in variant detection with reference laboratories. The assay’s reproducibility was confirmed with 100% inter-precision and intra-precision concordance. The RareVision WGS assay provides detailed genomic insights, enhancing the diagnosis and management of rare genetic disorders by offering a comprehensive and accurate genomic profiling tool.

## 1. Introduction

The clinical application of next-generation sequencing (NGS) is revolutionizing disease management by providing precise genetic insights for personalized therapy. Detecting causative variants is essential for diagnosing and managing rare diseases due to their genetic origins. Over 7,000 rare genetic diseases have been identified, many presenting in early childhood, making comprehensive genomic profiling crucial for understanding their complex etiologies [1-3]. Establishing a diagnosis is often complex, time-consuming, and costly, frequently resulting in misdiagnosis, ineffective treatments, and prolonged suffering for patients [4, 5].

Traditional diagnostic methods, such as chromosomal microarray analysis (CMA) for detecting copy number variations (CNVs) and targeted panel sequencing for single-nucleotide variants (SNVs) and small insertions and deletions (indels), are limited by their need for prior diagnostic hypotheses, reducing their clinical utility for rare genetic diseases. Whole-exome sequencing (WES), which focuses on protein-coding regions, has improved diagnostic yield [6-9]but has significant limitations, such as missing large indels, structural variations (SVs), absence of heterozygosity (AOH), short tandem repeats (STRs), mitochondrial variants, transposable elements (TEs), and intronic SNVs [6].

Whole Genome Sequencing (WGS) offers a comprehensive approach, surpassing traditional methods in scope and diagnostic yield [6]. Multiple independent studies have presented evidence for the clinical utility of WGS in the clinical diagnosis of rare genetic diseases [10-12]. This paper presents the analytical validation and clinical validation of a WGS-based germline comprehensive genomic profiling test. The study demonstrates the test’s precision, sensitivity, and reproducibility, confirming its capability to provide accurate and reliable genomic insights. These findings support WGS as a crucial tool in the genetic diagnostic landscape, potentially reducing the prolonged diagnostic process that many patients and their families endure.

## 2. Data and Methods

### 2.1 Test principle and intended use

WGS assay (RareVision -Inocras, San Diego, CA) is designed to detect causative germline variants in patients with an undiagnosed disease. This assay is adept at systematically identifying a wide range of genomic aberrations in germline specimens. SNVs, multiple nucleotide variants (MNVs), indels, CNVs, SVs and TEs. The intended use of WGS is to identify genomic alterations that have established and/or potential clinical relevance in the context of rare genetic diseases.

### 2.2. Patient and Reference Materials

In this study, a dual-methodology approach was implemented, combining well-characterized reference materials with patient-derived specimens for robust analysis. The reference materials included commercially available, well-characterized cell lines from Coriell Institute for Medical Research (Camden, NJ), SeraCare (Milford, MA), and Horizon Discovery (Cambridge, UK). These DNA specimens have known variants encompassing the following variant types: SNV, indel, CNV, and SV (see **Table 1** for a specific list of the cell lines). For assay precision and quality control, the NA12878 cell line from the Coriell Institute was utilized. This cell line is known for its extensively documented human genome reference, providing a reliable external quality control measure throughout the validation process.

**Table 1:** Reference material cell Lines for analytical validation.

| Description | Source |
| --- | --- |
| *KRAS* Reference Standard | Horizon Discovery |
| *BRCA* Reference Standard | Horizon Discovery |
| NA12878 | Coriell Institute for Medical Research |
| NA24143 | Coriell Institute for Medical Research |
| NA24385 | Coriell Institute for Medical Research |
| NA24631 | Coriell Institute for Medical Research |
| NA24694 | Coriell Institute for Medical Research |
| NA24695 | Coriell Institute for Medical Research |
| *BRCA1/2* LGR Inherited Mutation Mix | SeraCare |

The clinical validation of the WGS assay was conducted using a select cohort of 30 residual patient samples. These samples were obtained from a cohort registry study at Seoul National University (IRB-H-0905-041-281 and IRB-H-2202-045-1298). Selection was based on the availability of adequate residual genomic DNA extracted from blood and buccal swabs provided by Seoul National University, a CAP-accredited laboratory (CAP number: 6672401). The inclusion criteria for sample selection included a prior rare disease diagnosis. The study protocol was approved by the Seoul National University Institutional Review Board, ensuring compliance with the ethical standards outlined in the Declaration of Helsinki, and all participants provided written informed consent.

### 2.3 Regulatory standards

WGS is conducted at Inocras, a facility accredited by the College of American Pathologists (CAP) and certified under the Clinical Laboratory Improvement Amendments (CLIA) in the United States. CLIA provides federal regulatory standards that apply to all clinical laboratory testing performed on humans in the United States, excluding clinical trials and basic research.

### 2.4 Library Preparation

For this study, DNA samples were processed using the Watchmaker DNA Library Preparation Kit (Watchmaker Genomics, Inc., Boulder, CO). Initially, the DNA underwent enzymatic fragmentation, followed by size selection to achieve the desired insert size. This step was crucial for optimizing library construction. Unique dual index (UDI) adapters were then attached to each sample, facilitating downstream demultiplexing. The prepared libraries were quantified and normalized before pooling.

Key quality control measures included assessing the library size distribution targeting an average fragment size of approximately 300 bp, and quantifying yield, with a minimum threshold of 15 ng/uL. Library size was determined by the TapeStation 4200 System (Agilent Technologies, Santa Clara, CA). Concentration determinations were made using the Qubit 1x Broad Range dsDNA Assay Kit in conjunction with a Qubit 2.0 Fluorometer (ThermoFisher Scientific, Waltham, MA, USA). Prepared libraries were preserved at ≤ –20°C when not immediately processed to the capture stage, adhering to the kit manufacturer’s storage guidelines.

### 2.5 Sequencing and Data Analysis Pipeline

The comprehensive genomic analysis was performed using the WGS RareVision system (Inocras Inc., San Diego, CA, USA). The prepared DNA libraries were sequenced on the Illumina NovaSeq 6000 sequencing system (Illumina Inc.), which incorporates PhiX control for accuracy. The target average depth of coverage was 30x (deduplicated unique reads).

The raw sequence data generated were aligned to the human reference genome GRCh38 using the BWA-MEM algorithm. Post-alignment sequence analysis was conducted through a suite of validated bioinformatics tools integrated into a custom data-processing pipeline tailored for NGS platforms. This pipeline includes a comprehensive assessment of genetic variants, designed to evaluate the impact of each variant. For SNVs, MNVs, and indels, data from well-curated databases such as the 1000 Genomes Project [11], gnomAD [13], ExAC [14], TOGOVAR [15], and GASP[16], in addition to proprietary databases (GINS_PM_PON v1.4), were employed. SVs were analyzed using public resources like gnomAD SV v2.1.4.8 and internal databases.

#### The variant analysis protocol is structured as follows

⍰ Complete inclusion of the protein-coding regions.
⍰ Trio de novo analysis extends beyond protein coding to regulatory and intronic regions.
⍰ For clinically relevant genes, the analysis also covers known non-coding regions.
⍰ SV analysis prioritizes regions that include protein-coding sequences.
⍰ TE gene rearrangements focus on genes correlating with clinical symptoms.
⍰ Chromosomal copy number profile analysis is integrated into the assessment.

The detected SNVs, MNVs, and indels were categorized into five classes: pathogenic (P), likely pathogenic (LP), likely benign (LB), benign (B), and variants of uncertain significance (VUS), following the 2015 ACMG/AMP guidelines [17]. This study primarily addressed P and LP mutations, but also considered VUSs, particularly when rare variants in critical genes warranted further examination of clinical symptoms. Variant classification may be revised based on emerging research from relevant literature and databases. SVs and TEs involving exon regions were deemed pathogenic. CNVs were also classified into five categories (P, LP, B, LB, VUS) according to ACMG/ClinGen standards [18].

#### Results are grouped into four categories

⍰ Positive: A P/LP variant is identified in a gene associated with autosomal dominant/X-linked (AD/XL) clinically relevant disease, or two or more pathogenic/likely pathogenic variants are found in a gene for autosomal recessive (AR) disease.
⍰ Inconclusive: A VUS is identified in a gene associated with AD/XL clinically relevant disease, more than one VUS is identified in a gene responsible for AR disease, or only one P/LP variant is identified in a gene causing AR disease. Variants are found in genes of uncertain significance that match the patient’s clinical symptoms.
⍰ Negative: No clinically relevant variants are detected.
⍰ Secondary (Incidental): According to ACMG guidelines, if pathogenic variants are discovered in genes listed in Appendix I, these are reported as incidental findings, regardless of their association with the patient’s symptoms [19].

### 2.6 Orthogonal Testing

The clinical validation samples were previously sequenced by a CAP-accredited laboratory (Seoul National University Hospital) and therefore the known outcomes were utilized as the orthogonal method testing to ensure method comparison is acute. The known outcomes from the Seoul National University Hospital analysis were compared against the data generated from the clinical validation cohort.

## 3 Results

The RareVision WGS test was subjected to analytical validation, which included assessing various parameters such as nucleic acid extraction, sequencing accuracy, and data analysis precision. The assay’s performance was evaluated using a spectrum of cell lines and reference standards from established commercial sources, alongside clinical samples verified through orthogonal testing methods. WGS achieved a minimum of 20x sequencing depth (**Figure 1**).

**Figure 1:**
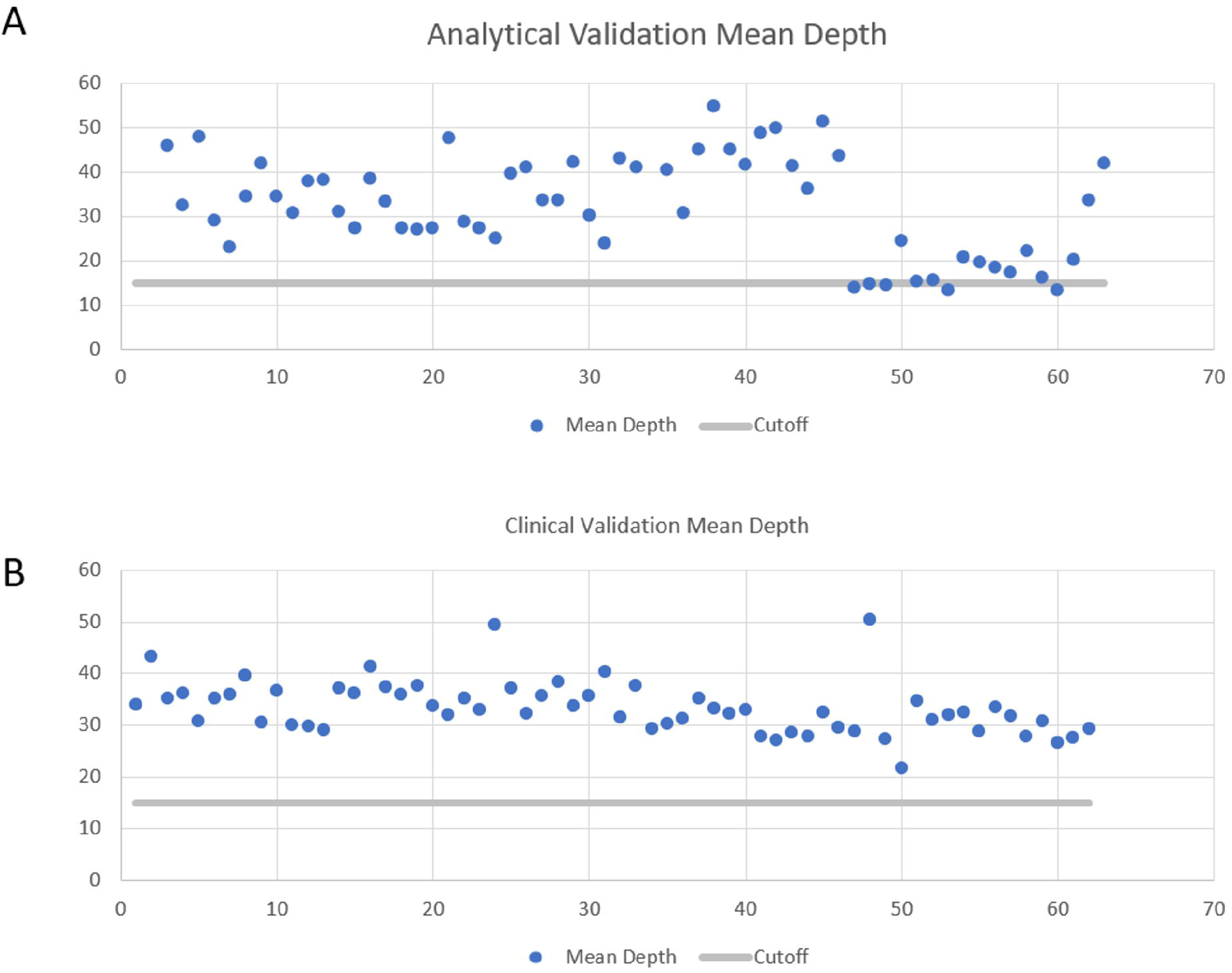
Mean Depth per Sample in Validation Runs. (A) Analytical Validation Samples and (B) Clinical Validation Samples. The assay’s performance was evaluated using a spectrum of cell lines and reference standards from established commercial sources, alongside clinical samples verified through orthogonal testing methods. Whole Genome Sequencing achieved a minimum of 20x sequencing depth across samples. The blue point represents the sample depth, while the grey line indicates the baseline depth of 15x.

### 3.1. Analytical Sensitivity and Positive Predictive Value for SNV and Indel Detection of Whole Genome Sequencing

A true-positive (TP) was a variant of interest (VOI) in the commercial cell lines identified by both the Genome in a Bottle (GIAB) consortium and the WGS assay. Conversely, a false-negative (FN) variant was recognized by the GIAB consortium but was missed by the WGS assay. The assay’s sensitivity was quantified using the formula: TP / (TP + FN) = Assay Sensitivity. This calculation, applied to data from six reference cell lines (GIAB), resulted in an analytical sensitivity of 99.4% for SNVs (95% CI: 99.14% - 99.72%) and 98.7% for indels (95% CI: 98.43%-99.05%; **Table 2**).

**Table 2:**
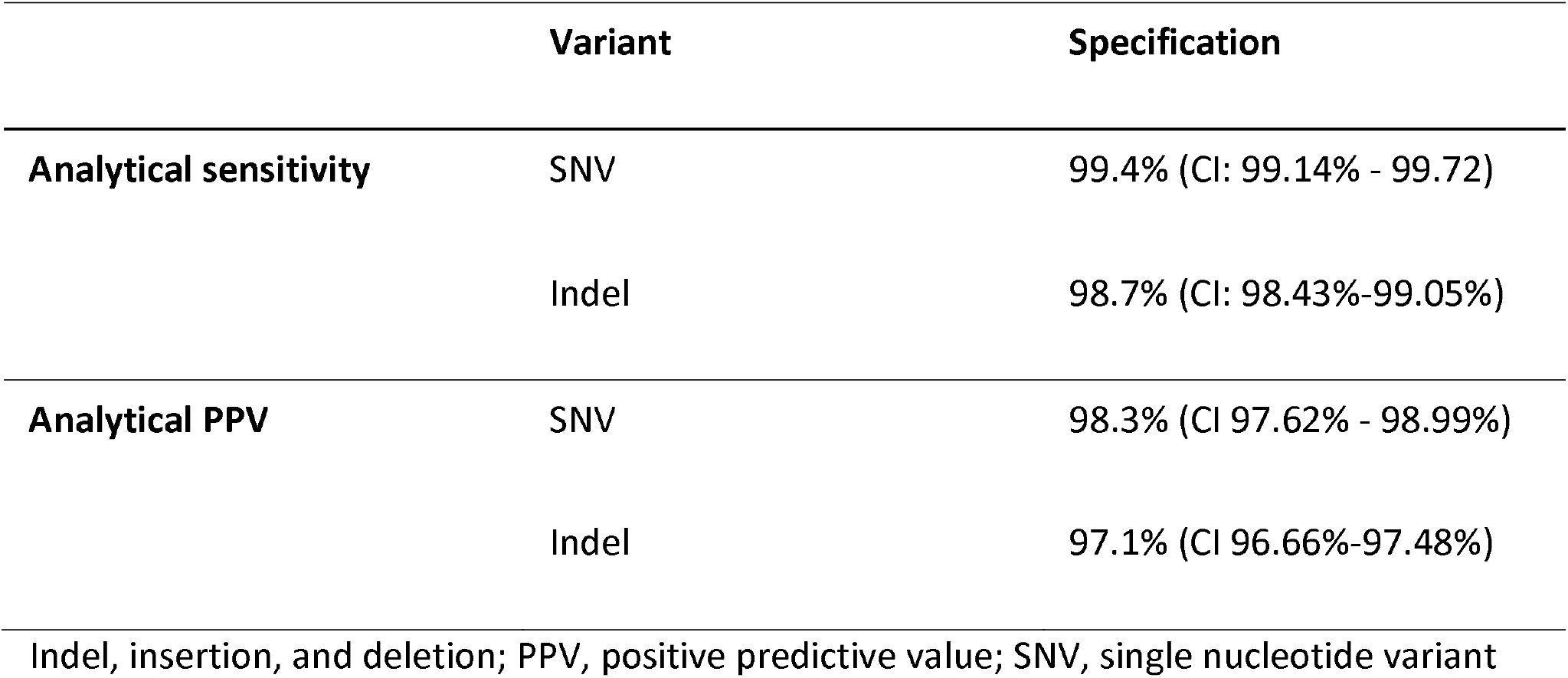
Overall performance of whole genome sequencing (WGS)

In this study, false positives (FP) were defined as VOI reported by the WGS assay at a locus that had been previously reported as a wild-type sequence in the GIAB database. To calculate the assay’s Positive Predictive Value (PPV), the formula: PPV = TP / (TP + FP) was used, where TP represents true positives. The WGS analytical PPV, evaluated against the GIAB standard revealed a PPV of 98.3% for SNVs (95% Confidence Interval [CI]: 97.62% - 98.99%) and 97.1% for indels (95% CI: 96.66%-97.48%), as detailed in **Table 2**.

### 3.2. Limit of Detection (LOD)

Variant Allele Frequency (VAF): VAF is not a reportable value for the WGS assay, as germline variants are represented in either a homozygous or heterozygous manner.

Sample Input: The amount of DNA used for library preparation in the WGS assay followed the manufacturer’s recommendations, ranging from 1 to 500 ng. Previous internal feasibility studies have indicated that input values below 50 ng do not produce adequate amplification for complete sequencing. Consequently, for this validation study, 50 ng was established as the lowest sample input value. Cell line samples from the GIAB were processed through library preparation at varying amounts of DNA input to verify that there was no correlation between the amount of input and the sequencing metrics. SNV sensitivity, SNV PPV, indel sensitivity, and indel PPV were calculated against the GIAB’s reference standard. The variations in library preparation input demonstrated no significant effect on the sequencing metrics, as depicted in **Figure 2**.

**Figure 2:**
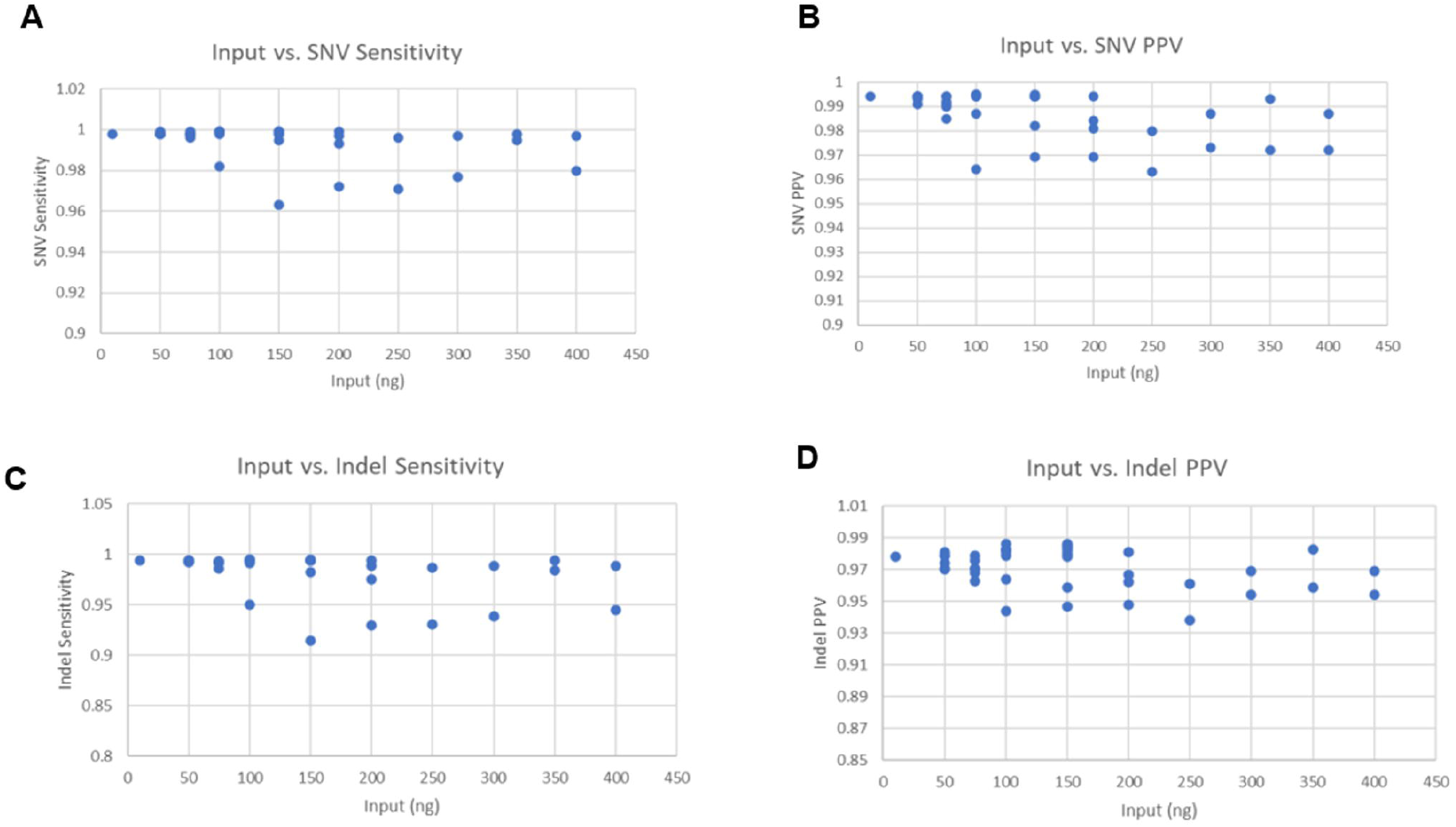
Impact of DNA Input on Sequencing Metrics. Cell line samples from the Genome in a Bottle (GIAB) were processed through library preparation at varying DNA input levels to verify the lack of correlation between the input amount and sequencing metrics. Sensitivity and positive predictive value (PPV) were calculated against the GIAB reference standard. The results indicate that variations in library preparation input have no significant effect on sequencing metrics, validating 50 ng as the lowest sample input value. (A) Sensitivity of single nucleotide variants (SNVs) across varying DNA input amounts. (B) PPV of SNVs across varying DNA input amounts. (C) Sensitivity of insertions/deletions (indels) across varying DNA input amounts. (D) PPV of indels across varying DNA input amounts.

### 3.3. Reproducibility

#### Inter-Precision Reproducibility

Inter-precision reproducibility was assessed by calculating concordance between two operators on the same set of samples on different days in the laboratory. Two trained clinical laboratory scientists (CLS), Operator 1 and Operator 2, each performed library preparation and sequencing on the 30 unique clinical validation patient samples. The operators performed their bench work on different days and used different sides of the sequencer (side A and side B) on a NovaSeq 6000, which is the sole sequencer used at Inocras. The inter-precision reproducibility was found to be 100%, with both operators’ sample sets receiving 100% concordant calls.

#### Intra-Precision Reproducibility

Intra-precision reproducibility was assessed by calculating concordance within operators by running duplicates of the same specimen DNA. One trained CLS performed library preparation and sequencing in duplicate on 30 DNA specimens. These samples were run in analytical validation runs 1 and 2. Both analytical validation flow cells received concordant calls.

### 3.4. Clinical Validation

Clinical validation of the assay was performed by analyzing 30 samples that had previously been sequenced by Seoul National University Hospital, a CAP-accredited laboratory. This existing data served as the orthogonal comparator for our study. The analysis confirmed that every variant detected by Seoul National University Hospital was also identified by RareVision. Each of the 30 clinical validation samples contained detectable variants, demonstrating the intra-assay precision of the method, which was determined to be 100% (30/30).

## 4. Discussion

In the rapidly advancing field of precision medicine, the role of comprehensive genomic profiling in diagnosing and managing rare genetic diseases is increasingly recognized. The RareVision WGS assay has been carefully developed and validated to meet the needs of this paradigm by enabling a thorough examination of genomic variations that underlie rare genetic diseases. This assay has undergone extensive analytical and clinical validation, proving to be a highly reliable and accurate tool for comprehensive genomic profiling. The RareVision WGS assay exhibits high sensitivity and specificity for SNVs, indels, and SVs. The analytical validation results demonstrated a sensitivity of 99.4% for SNVs and 98.7% for indels, with corresponding PPVs of % and 98.7%, respectively. These metrics underscore the assay’s robustness and accuracy in detecting a wide array of genomic variants, essential for the precise diagnosis of rare genetic disorders.

The clinical validation of the RareVision WGS assay involved a cohort of 30 samples previously sequenced by a CAP-accredited laboratory. The complete concordance between the variants identified by RareVision and those detected by the reference laboratory highlights the assay’s high accuracy and reliability. This validation process is crucial for establishing the assay’s clinical utility and ensuring that it can deliver consistent and dependable results in real-world settings.

The comprehensive nature of WGS allows for the detection of a broad spectrum of genomic aberrations, including SNVs, indels, CNVs, SVs, and TEs. This capability positions WGS as a superior alternative to traditional diagnostic methods such as CMA and targeted panel sequencing, which are limited in scope and diagnostic yield. The ability to capture a more complete genomic profile can significantly shorten the diagnostic journey for patients with rare diseases, enabling more timely and accurate diagnoses.

The findings from this study strongly support the broader adoption of WGS in clinical practice, particularly for diagnosing rare genetic disorders. The high precision, sensitivity, and reproducibility of the RareVision WGS assay, combined with its comprehensive genomic coverage, make it a valuable tool in the genetic diagnostic landscape. By facilitating early and accurate diagnosis, WGS has the potential to improve patient outcomes and reduce the healthcare burden associated with prolonged diagnostic processes.

## 5. Conclusion

In conclusion, the RareVision WGS assay offers a robust and comprehensive solution for genomic profiling, making it a valuable tool for the diagnosis and management of rare genetic diseases.

## Data Availability

The patients participating in this study did not consent to the public release of sequencing data. The WGS pipeline and associated algorithms are proprietary to Inocras Inc.

## Acknowledgments

The authors would like to thank all patients and our collaborators at Seoul National University Hospital, Seoul National University College of Medicine.

## Ethics approval and consent to participate

This work combined data from the parent study (IRB-H-0905-041-281 and IRB-H-2202-045-1298) that had previously obtained informed consent from participants and IRB approval. The authors state that they have followed the principles outlined in the Declaration of Helsinki.

## Consent for Publication

In alignment with ethical guidelines, written informed consent for the publication of their data and/or images was obtained from all participants involved in the study. This consent was part of the initial ethical approval process (IRB-H-0905-041-281 and IRB-H-2202-045-1298). Additionally, all patient information presented in this publication has been anonymized to protect patient privacy and confidentiality.

